# An Immunobridging study to evaluate the neutralizing antibody titer in adults immunized with two doses of either ChAdOx1-nCov-19 (AstraZeneca) or MVC-COV1901

**DOI:** 10.1101/2022.02.26.22271364

**Authors:** Josue Antonio Estrada, Chien-Yu Cheng, Shin-Yen Ku, Hui-Chun Hu, Hsiu-Wen Yeh, Yi-Chun Lin, Cheng-Pin Chen, Shu-Hsing Cheng, I-Feng Lin

**Author notes:** **Corresponding author:** Chien-Yu Cheng, Division of Infectious Diseases, Department of Internal Medicine, Taoyuan General Hospital, Ministry of Health and Welfare, No. 1492, Zhongshan Rd., Taoyuan Dist., Taoyuan City 330, Taiwan (R.O.C.). **Alternative corresponding author:** I-Feng Lin Prof. Institute of Public Health, School of Medicine National Yang-Ming Chiao Tung University, 155 Li-Nong Street, Pei-Tou, Taipei, Taiwan R.O.C <u>Tel:</u>, <u>E-mail:</u>.

## Abstract

**Background:** Rapid development and deployment of vaccine is crucial to control the continuously evolving COVID-19 pandemic. Placebo-controlled phase 3 efficacy trial is still standard for authorizing vaccines in majority of the world. However, due to lack of cases or participants in parts of the world, this has not always been feasible. An alternative to efficacy trial is immunobridging, in which the immune response or correlates of protection of a vaccine candidate is compared against an approved vaccine. Here we describe a case study where our candidate vaccine, MVC-COV1901, has been granted for emergency use authorization (EUA) locally based on the non-inferiority immunobridging process.

**Methods:** The per protocol immunogenicity (PPI) subset from the MVC-COV1901 phase 2 trial was used for this study and consisted of 903 subjects who have received two doses of MVC-COV1901 as scheduled in the clinical trial. The comparator set of population consisted of 200 subjects of ≥ 20 years of age who were generally healthy and have received two doses of AstraZeneca ChAdOx nCOV-19 (AZD1222) recruited from Taoyuan General Hospital, Ministry of Health and Welfare.

**Results:** MVC-COV1901 was shown to have a geometric mean titer (GMT) ratio lower bound 95% confidence interval (CI) of 3.4 against the comparator vaccine and a seroconversion rate of 95.5% at the 95% CI lower bound, which both exceeded the criteria set by the Taiwan regulatory authority for EUA approval. These results supported the EUA approval of MVC-COV1901 by the Taiwanese regulatory authority in July 2021. Following the consensus of the International Coalition of Medicines Regulatory Authorities (ICMRA), countries from the Access Consortium has recently adopted the use of immunobridging studies as acceptable for authorizing COVID-19 vaccines in lieu of efficacy data.

**Conclusion:** The data presented in the study showed that it is reasonably likely that the vaccine efficacy of MVC-COV1901 is similar or superior to that of AZ. Data could be used in support of further vaccine development and regulatory approval.

## Introduction

MVC-COV1901 is a protein subunit SARS-CoV-2 vaccine based on the stabilized prefusion spike protein S-2P adjuvanted with CpG 1080 [1]. From 2020 to early 2021, Taiwan has been spared from the worst of the pandemic, which recorded only a total of local and imported of 525 cases in 2020 and 339 cases in 2021 prior to the local outbreak in May 2021 [2]. As a result, it was not feasible to conduct placebo-controlled efficacy trial locally in Taiwan at the time. In response, Taiwan health authorities designed a pathway to EUA for all local vaccine candidates based on immunobridging, which compares the immune response of a vaccine candidate with an approved vaccine for comparison [3]. Over 3,800 participants were enrolled in a phase 2 clinical trial for MVC-COV1901 and the results of this large-scale phase 2 trial allowed the Taiwan regulatory authorities to examine the safety and immunogenicity to the comparator vaccine which has already been approved in Taiwan for non-inferiority [3, 4]. MVC-COV1901 was approved by the Taiwan Food and Drug Administration (TFDA) in July 2021, making it among the first COVID-19 vaccine approved using immunobridging study prior to the availability of efficacy data [5]. In June 2021, experts from regulatory authorities around the world convened at a workshop for the future of COVID-19 vaccine development, and consensus was reached for the use of well-justified and appropriately designed immunobridging studies in place of clinical endpoint efficacy studies when they are not feasible [6]. In September 2021, the consensus position has since been taken up by the Access Consortium, which consisted of regulatory authorities from the UK, Australia, Canada, Singapore and Switzerland, to accept immunobridging studies as sufficient for authorizing COVID-19 vaccines in these countries [7, 8]. This manuscript thus illustrates an example of a COVID-19 vaccine approved with immunobridging study, which has gradually become recognized by health regulatory authorities worldwide.

## Methods

### Clinical trial and sample population

The MVC-COV1901 phase 2 trial was a prospective, randomized, double-blind, placebo-controlled, and multicenter study to evaluate the safety, tolerability, and immunogenicity of the SARS-CoV-2 vaccine candidate MVC-COV1901 (NCT04695652) [4]. The main study of the trial consisted of 3,844 subjects of ≥ 20 years of age who were generally healthy or with stable pre-existing medical conditions recruited from eleven sites in Taiwan, and this population was used for safety analysis [4]. The per protocol immunogenicity (PPI) subset from the MVC-COV1901 phase 2 trial was used for this study and consisted of 903 subjects who have received two doses of MVC-COV1901 as scheduled in the clinical trial. The comparator set of population consisted of 200 subjects of ≥ 20 years of age who were generally healthy and have received two doses of AstraZeneca ChAdOx nCOV-19 (AZD1222) recruited from Taoyuan General Hospital, Ministry of Health and Welfare.

The trial protocol and informed consent form were approved by the Taiwan Food and Drug Administration (FDA) and the ethics committees at the participating sites. The institutional review boards included the Chang Gung Medical Foundation, National Taiwan University Hospital, Taipei Veterans General Hospital, Tri-Service General Hospital, Taipei Medical University Hospital, Taipei Municipal Wanfang Hospital, Taoyuan General Hospital Ministry of Health and Welfare, China Medical University Hospital, Changhua Christian Hospital, National Cheng Kung University Hospital, and Kaoshiung Medical University Hospital. The trial was done in accordance with the principles of the Declaration of Helsinki and good clinical practice guidelines.

### Vaccines

Medigen COVID-19 vaccine or MVC-COV1901, a subunit vaccine consisting of the prefusion spike protein (S-2P) adjuvanted with 750 μg CpG 1018 and 375 μg aluminum hydroxide. A standard 0.5 mL dose contains 15 µg of the Spike-2P. Both are delivered instramuscularly at the deltoid region. The comparator vaccine is ChAdOxAZD1222, an adenoviral vector vaccine developed by Oxford University and AstraZeneca served at multi-dose vials. Each dose of vaccine is 0.5 mL and contains 5×10^10^ viral particles.

### Immunobridging study

According to the TFDA document, the follow criteria were set for a candidate vaccine to be granted EUA in Taiwan [3]:

- Clinical data: Immunobridging study to evaluate the immunogenicity of locally developed vaccine against a comparator vaccine which has already been approved in Taiwan.
- Safety data: At least 3,000 subjects were required to be tracked for at least one month for safety data after the last dose and all subjects to be followed for a median of two months after the last dose.

As the AstraZeneca ChAdOx1 nCoV-19 (AZD1222) vaccine was the first COVID-19 vaccine to be approved in Taiwan, it was chosen as the comparator vaccine for which the locally developed vaccines are to be benchmarked with [3]. The immunobridging criteria were to fulfill the following endpoints for serum samples 28 days after the second dose (Day 57) in population under the age of 65 [3, 9]:

1. The lower limit of the 95% confidence interval of the geometric mean titer ratio (GMTR) of the prototype strain live virus neutralizing antibodies for the MVC-COVID19 vaccine group to the external control group must be greater than 0.67, as shown in a blood test 28 days after the second dose;
2. The sero-response level was defined as the neutralizing antibody titers against the prototype strain live virus at 28 days after receiving the second dose of the vaccine, at the referred point of 60% reverse accumulative distribution curve for external control group The lower limit of the 95% confidence interval for the sero-response rate (the proportion of subjects whose neutralizing antibody titers against the prototype strain live virus, at 28 days after receiving the second dose of the MVC-COVID19 vaccine are above the sero-response level) must be greater than 50%

### Live SARSDCoVD2 neutralization assay

Neutralizing antibody titers against the Wuhan prototype SAR-CoV-2 were determined by SARS-CoV-2 live virus neutralization assay as described previously [4].

### Statistical analysis

Descriptive statistics were obtained and presented for the population’s characteristics. GMTs were estimated from neutralizing antibody titers measured at 28 days after the second dose of the study intervention. Geometric mean titer ratio (GMTR) is calculated as the GMT of MVC group over the GMT of AZ group.

The GMTs are presented with their two-sided 95% CI. To assess the magnitudes of differences in immune response between the two vaccines, an analysis of covariance (ANCOVA) model was used. The model included the log-transformed antibody titers at Day 57 as the dependent variable, vaccine group (AZD1222 and MVC-COV1901) as an explanatory variable and adjusted for age, BMI, gender and comorbidity profile. The 95% CI for the adjusted neutralizing antibody titers of each vaccine group were obtained. Then adjusted GMT and corresponding 95% CI were back-transformed to the original scale. A worst modified subset of the sample was created which included only GMT values ≤ 67th percentile at Day 57. Reverse cumulative distribution (RCD) curves were constructed for data 28 days after the 2^nd^ dose for the AZD1222 and MVC-COV1901 groups.

## Results

### Demographics

The mean age of AZD1222 (AZ) and MVC-COV1901 (MVC) groups were similar at 42.9 and 45.7 years, respectively (Table 1). However, only six (3%) subjects were over age of 65 in the AZ group compared to 221 (24.4%) in MVC-COV1901, though this is due to the study design of the MVC-COV1901 trial to enroll at least 20% of participants to be over age of 65 [4]. In terms of gender, the AZ group had higher percentage of females (59.8%) than that of MVC group (42.3%). Other discrepancies between the two groups include the higher proportion of comorbidity in the AZ group (38.1%) compared to the MVC group (19.3%). In order to investigate the effects of these factors for the immunobridging process, we performed sensitivity analyses based on age group, gender, and comorbidity.

**Table 1.** Demographics of the population groups.

| Item\MVC lot | <65 years |  | All ages |  |
| --- | --- | --- | --- | --- |
|  | AstraZeneca |  | AstraZeneca |  |
|  | ChAdOx1 | MVC-COV1901 | ChAdOx1 | MVC- |
|  | nCoV-19 | PPI subset | nCoV-19 | COV1901 |
|  | N = 194 | N=682 | N = 200 | N=903 |
| **Age (years)** |  |  |  |  |
| Mean (SD) | 42.2 (11.1) | 38.02 (11.03) | 42.9 (11.7) | 45.68 (16.64) |
| Median (IQR) | 41(16.75) | 37.0 (17.0) | 41.5(17.2) | 42.0 (32.0) |
| Min~Max | 22.0~64.0 | 20.0~64.0 | 22.0~69.0 | 20.0~ 87.0 |
| **Gender** |  |  |  |  |
| Male | 78 (40.2) | 386 (56.6) | 80 (40.0) | 521 (57.7) |
| Female | 116 (59.8) | 296 (43.4) | 120 (60.0) | 382 (42.3) |
| **BMI (kg/m<sup>2</sup>)** |  |  |  |  |
| Mean (SD) | 25.3 (4.5) | 24.9 (4.3) | 25.2(4.6) | 24.9 (4.1) |
| Median (IQR) | 254.6 (5.97) | 24.3 (5.7) | 24.5(5.97) | 24.4 (5.3) |
| Min~Max | 16.6~39.6 | 14.3~45.2 | 17.01~37.5 | 17.7 ~36.9 |
| **BMI group** |  |  |  |  |
| <30 kg/m<sup>2</sup> | 159 (81.96) | 594 (87.1) | 798 (88.4) | 164 (82.0) |
| >=30 kg/m<sup>2</sup> | 35 (18.04) | 88 (12.9) | 105 (11.6) | 36 (18.0) |
| **Pre-vaccination neutralizing antibody status** |  |  |  |  |
| Seropositive | 2 (1.03) | 8 (1.17) | 2 (1.0) | 10 (1.11) |
| Seronegative | 192 (98.97) | 674 (98.83) | 198 (99.0) | 893 (98.9) |
| **Comorbidity Category** |  |  |  |  |
| at least one comorbidity | 74 (38.1) | 89 (13.0) | 123 (61.5) | 729 (80.7) |
| No comorbidity | 120 (61.9) | 593 (87.0) | 77 (38.5) | 174 (19.3) |

### Immunogenicity

In subjects of under age of 64, at 28 days after the second dose, the AZ and MVC groups had GMTs of 186 and 733, respectively (Figure 1). When including the above 65 years of age into the GMT calculation, the GMTs decreased to 184 and 662 for the AZ and MVC groups, respectively, due to the inclusion of lower neutralizing antibody titer levels in the older age group (Figure 1). According to the analysis for immunobridging study, the results showed that the lower limit of the 95% CI for the GMTR of the prototype strain live virus neutralizing antibodies between MVC and AZ groups was 3.4 times, which was greater than the requirement of 0.67 times. The lower limit of the 95% confidence interval for the sero-response rate of the MVC group was 95.5%, which was greater than the requirement of 50%.

**Figure 1.**
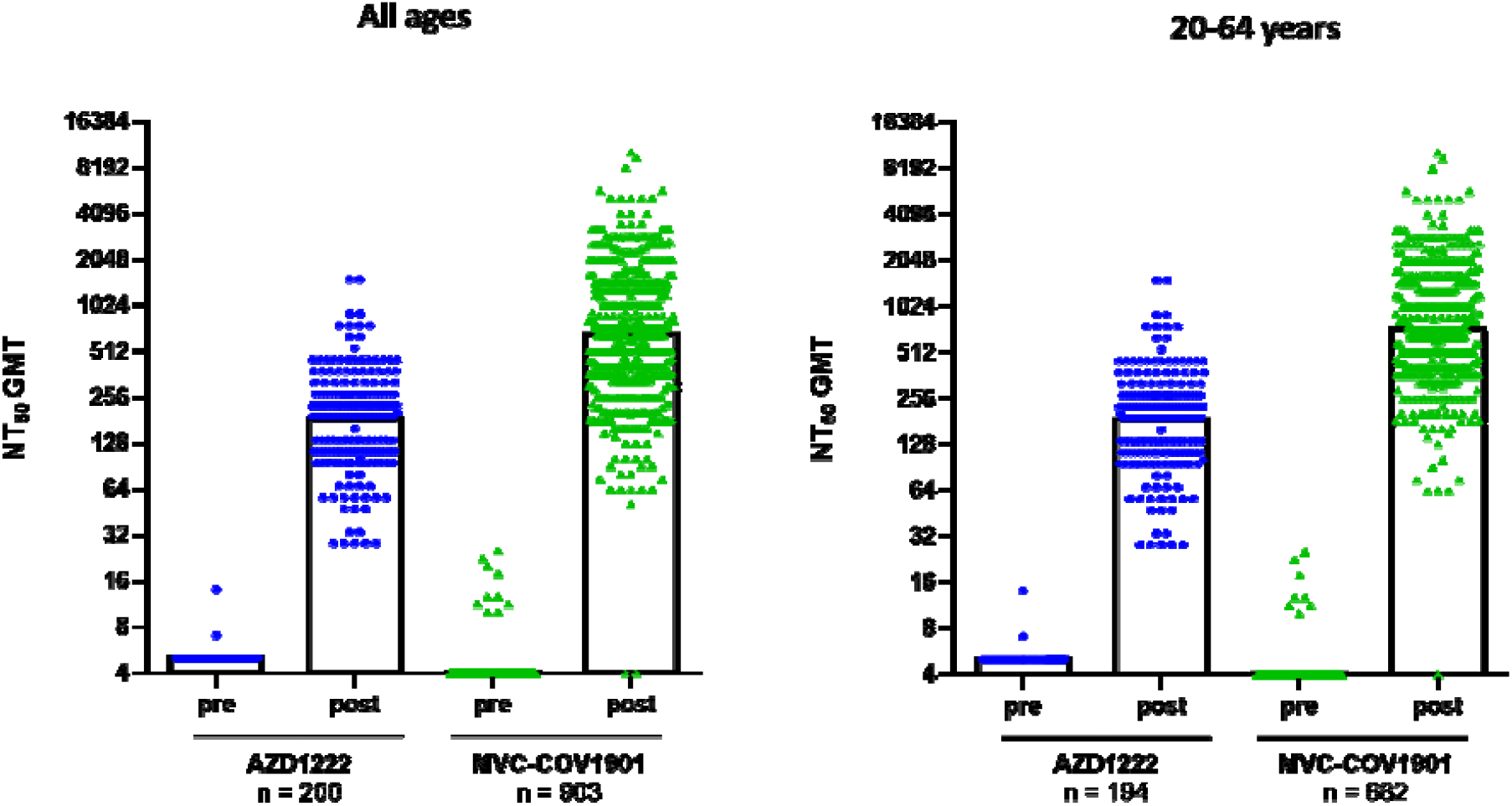
Neutralizing antibody titer in subjects immunized with two doses of either AZD1222 or MVC-COV1901 in all ages (left) and ages 20-64 years (right) Serum samples were taken before the first vaccination (pre) or 28 days (post) after the second dose of either vaccine and were subjected to live SARS-CoV-2 neutralization assay. The results are shown as 50% neutralizing titer (NT_50_) with symbols indicating individual NT_50_ values and the bars indicating the GMT of each group.

Illustrated in Figure 2 are the reverse cumulative distribution (RCD) curves of neutralizing antibody titers. Higher neutralizing antibody titers were observed in the MVC group. At the referred point of 60% for the AZ group, participants had neutralizing antibody titers equal to or less than 199.5 IU/mL. This is equivalent to approximately 90% of individuals given MVC.

**Figure 2.**
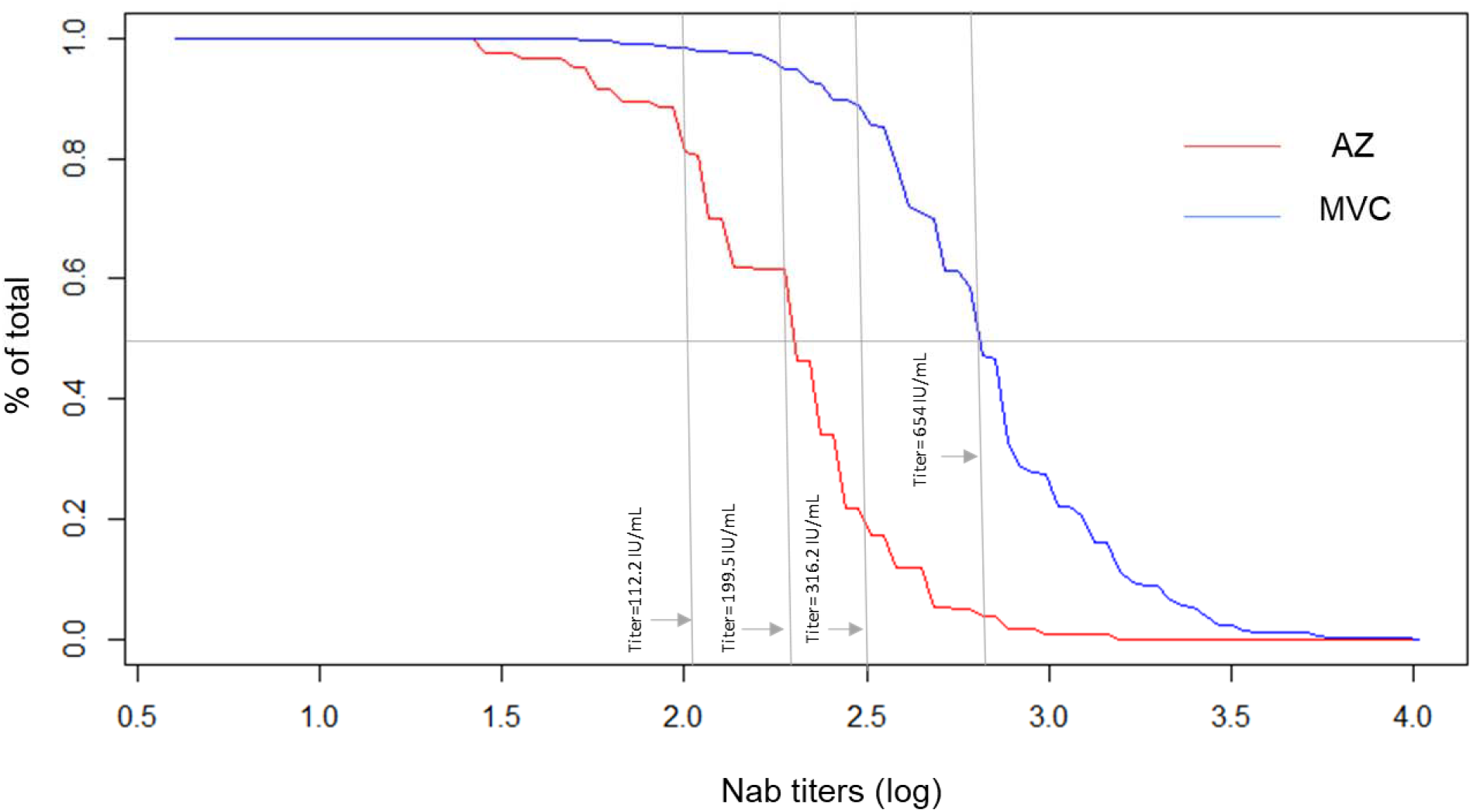
Reverse Cumulative Distribution Curve using log-transformed titers by percent of subjects who had neutralizing antibody titers 28 days following the second dose of AZ and MVC

### Sensitivity Analysis

The sensitivity analyses conducted to detect the robustness of GMT results reveal that both AZD1222 and MVC-COV1901 enhanced neutralizing antibody titers in both the subset of the younger individuals (aged 20-64 years) and the overall sample which includes older participants aged 65 years and above. The GMT Ratio (AZ vs. MVC) in the younger group which was 3.89 (95% CI: 3.45, 4.4) was comparable to the overall GMT Ratio of 3.55 (3.2, 3.97). The subgroup of younger individuals had higher GMTs for both AZ (185.97; 95% CI: 167.3, 206.7) and MVC (723.6; 95% CI: 683.7, 765.8) as compared to the overall GMTs for AZ (184.05; 95%CI: 166.5, 204.7) and MVC (654.07; 95% CI: 620.9, 689.03). Adjusting for age, sex, Body Mass Index (BMI) and comorbidity status, the population including 65 years and above has a more robust MVC response albeit comparable to those in the younger age group. The GMT ratio (MVC vs. AZ) in the overall sample is 3.80 (95% CI: 3.4, 4.3) while that of the younger subset is 3.78 (95% CI: 3.3, 4.3). Adjusted GMTs also indicate a better response among the younger age group (Table 2). Findings are consistent when looking at the worst modified subset of the sample (i.e. Day 57 titer ≤ 67th percentile). GMTs are lower in the overall sample which includes those 65 years and above, than in the younger group (Table 3). Adjusted GMT ratio is also comparable between the overall sample (2.70; 95% CI: 2.4,3.01) and the younger age group (2.69; 95% CI: 2.4, 3.02). Subgroup analyses based on gender and comorbidity profile show consistency in estimates across subgroups.

**Table 2.**
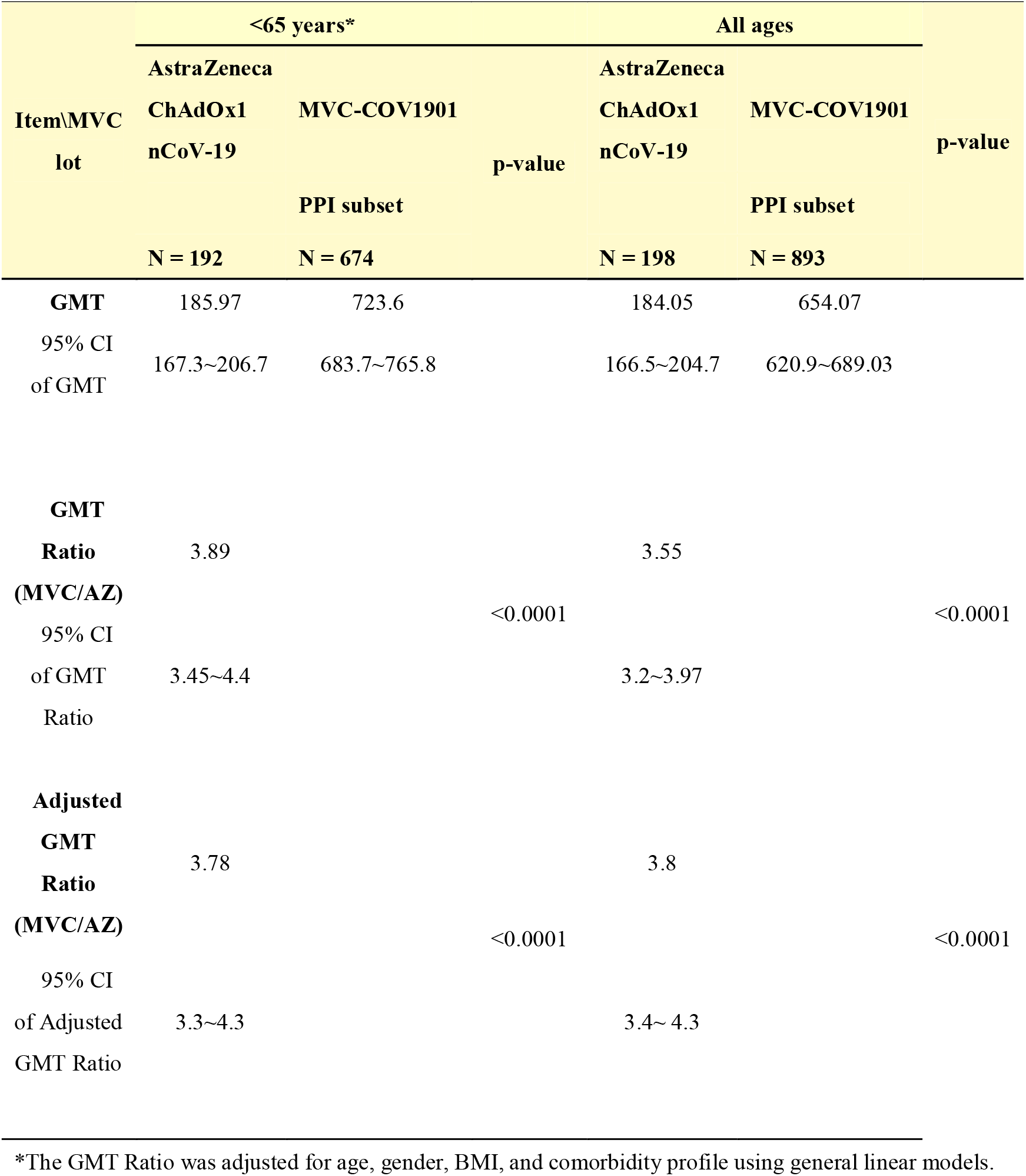
Neutralizing antibody titer in subjects immunized with either two doses of AZD1222 or MVC-COV1901 in all ages and ages 20-64 years at Day 57 (28 days after the second dose)

**Table 3.**
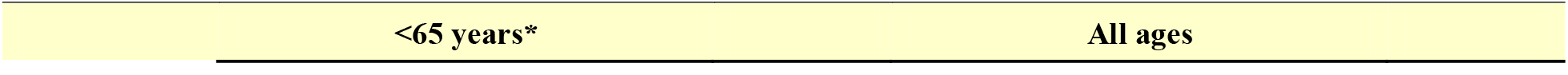
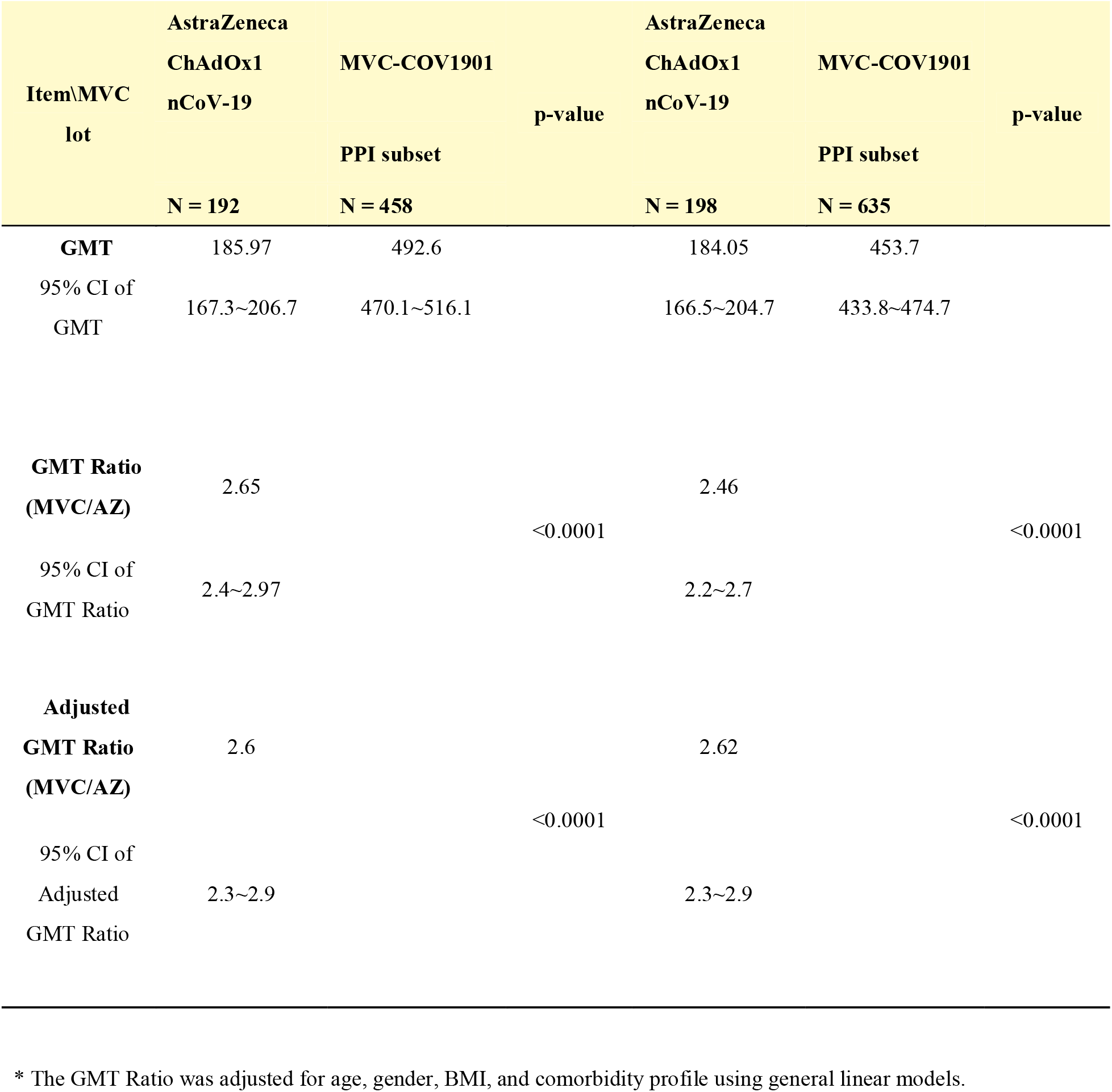
Worst modified PPI subset of neutralizing antibody titer in subjects immunized with either two doses of AZD1222 or MVC-COV1901 in all ages and ages 20-64 years at Day 57.

| Item\MVC<br>lot | AstraZeneca |  | p-value | AstraZeneca |  | p-value |
| --- | --- | --- | --- | --- | --- | --- |
|  | ChAdOx1 | MVC-COV1901 |  | ChAdOx1 | MVC-COV1901 |  |
|  | nCoV-19 | PPI subset |  | nCoV-19 | PPI subset |  |
|  | N = 192 | N = 458 |  | N = 198 | N = 635 |  |
| **GMT** | 185.97 | 492.6 |  | 184.05 | 453.7 |  |
| 95% CI of<br>GMT | 167.3~206.7 | 470.1~516.1 |  | 166.5~204.7 | 433.8~474.7 |  |
| **GMT Ratio**<br>**(MVC/AZ)** | 2.65 |  |  | 2.46 |  |  |
|  |  |  | <0.0001 |  |  | <0.0001 |
| 95% CI of<br>GMT Ratio | 2.4~2.97 |  |  | 2.2~2.7 |  |  |
| **Adjusted**<br>**GMT Ratio**<br>**(MVC/AZ)** | 2.6 |  |  | 2.62 |  |  |
|  |  |  | <0.0001 |  |  | <0.0001 |
| 95% CI of<br>Adjusted<br>GMT Ratio | 2.3~2.9 |  |  | 2.3~2.9 |  |  |

\* The GMT Ratio was adjusted for age, gender, BMI, and comorbidity profile using general linear models.

## Discussion

The major regulators considered that the evidence from studies with authorized COVID-19 vaccines is sufficient to support using neutralizing antibody titers as primary endpoint in an immunogenicity bridging study for authoring new COVID-19 vaccines. The decision had become relevant when clinical endpoint efficacy studies are less feasible. Our study followed the statement made by The International Coalition of Medicines Regulatory Authorities (ICMRA) [10] demonstrating the superiority of immunogenicity of MVC-COV1901 against AZ to predict vaccine effectiveness.

The regulator’s consortium recommended that the study design should be non-inferiority when the choice of the active comparator has demonstrated high efficacy, or superiority if the active comparator has modest efficacy. AZ being a product that has modest efficacy justified the need for demonstrating the superiority to gain regulatory approval. Our study tried to simulate the immunogenicity comparison between MVC-COV1901 and AZ done by the regulator in Taiwan through which a EUA was granted to the former vaccine. The comparative study was not done in a randomized, blinded study, but an external comparison. To increase confidence from the regulator’s perspective, a series of sensitivity analyses were conducted. The data showed that after omitting the highest 33 percent data points of neutralizing antibody titers, the results can still hold.

The neutralizing antibody titers were determined using World Health Organization (WHO)-certified reference standards, International Unit, IU/mL. The use of the standardized unit to report humoral immunogenicity could facilitate future cross-platform or cross-lab comparison. The study participants of both vaccines are mainly Taiwanese, reporting in IU/mL could follow cross-ethnicity comparison.

There are a few limitations of this study, first, the design is not randomized, double blinded, but an external comparison, which will compromise the level of evidence. Second, the cell-medicated immunity was not included in the comparative immunogenicity profile. Third, other characterizations that are of interests, including the waning immunity of both vaccines, the cross-reactivity against Variants of Concern (VoCs), were not explored.

The data presented in the study showed that it is reasonably likely that the vaccine efficacy of MVC-COV1901 is similar or superior to that of AZ. The data could be used in support of further vaccine development and regulatory approval.

## Data Availability

All data produced in the study are available upon reasonable request to the authors

## Acknowledgements

The study was funded by Medigen Vaccine Biologics (study sponsor) and the Taiwan Centres for Disease Control, Ministry of Health and Welfare.

## Author Contributions

Conceptualization, C.-Y. C. and I.-F. L.; methodology, I.-F. L.; software, C.-Y. C. and I.-F. L.; validation, S.-Y. K., C.-P. C., and Y.-C. L.; formal analysis, C.-Y. C., I.-F. L., and J.A.G.E.; investigation, C.-Y. C. and S.-H. C.; resources, C.-Y. C., H.-C. H., H.-W. Y. and S.-H. C.; data curation, I.-F. L. and J.A.G.E.; writing—original draft preparation, C.-Y. C. and J.A.G.E.; writing—review and editing, I.-F. L.; visualization, J.A.G.E and I.-F. L.; supervision, I.-F. L.; project administration, C.-Y. C. All authors have read and agreed to the published version of the manuscript.

## Competing Interests

J.A.G.E. is an employee of Medigen Vaccine Biologics (Taipei, Taiwan) and he has received grants from Taiwan Centres for Disease Control, Ministry of Health and Welfare, during the conduct of the study. All other authors declare no conflict of interest.

## Notes

### Clinical Trial

NCT04695652

### Author Declarations

The trial protocol and informed consent form were approved by the Taiwan Food and Drug Administration (FDA) and the ethics committees at the participating sites. The institutional review boards involved were the Chang Gung Medical Foundation, National Taiwan University Hospital, Taipei Veterans General Hospital, Tri-Service General Hospital, Taipei Medical University Hospital, Taipei Municipal Wanfang Hospital, Taoyuan General Hospital Ministry of Health and Welfare, China Medical University Hospital, Changhua Christian Hospital, National Cheng Kung University Hospital, and Kaoshiung Medical University Hospital. The trial was done in accordance with the principles of the Declaration of Helsinki and good clinical practice guidelines.

